# Automated Brain and CSF Volume Assessment in Infant Hydrocephalus Using Deep Learning

**DOI:** 10.64898/2026.05.07.26352592

**Authors:** Mingzhao Yu, Marcia Harumy Yoshikawa, Ariadna Sosa Luviano, Steven J. Schiff, Vishal Monga, Benjamin C. Warf, P. Ellen Grant, Jason Sutin, Pei-Yi Lin

## Abstract

Accurate brain and cerebrospinal fluid (CSF) volume assessment is essential for pediatric hydrocephalus management. Current clinical practice relies on linear measurements that fail to capture complex three-dimensional ventricular morphology, while quantitative volumetric assessment remains limited by laborious processing and lack of clinically optimized automated tools. This study developed a rapid, automated AI-based intracranial segmentation model suitable for clinical workflows. We retrospectively analyzed 167 T2-weighted MRI scans from infants with hydrocephalus, randomly split into training (60%), validation (20%), and hold-out test (20%) sets. All scans were manually segmented into CSF, brain parenchyma, and background. Our model integrates DenseNet and U-Net architectures with feature smoothness regularization to enhance generalizability. Performance was evaluated using Dice scores and absolute relative volume error (ARVE) compared with state-of-the-art methods. The AI model achieved Dice scores of 95.7% for CSF and 96.4% for brain parenchyma on the hold-out test set, significantly outperforming FSL FAST (85.0% and 77.9%) and contemporary deep learning approaches (90.4% and 89.7%). Processing time was 0.8 seconds per scan using GPU acceleration. The model demonstrated consistent performance across different hydrocephalus etiologies and effectively handled challenging scenarios including noise, artifacts, and variable resolution. This study successfully developed a robust MRI segmentation model demonstrating superior accuracy and efficiency compared to existing methods. By incorporating domain-specific enhancements, the model enables rapid, clinically viable brain and CSF volume estimation for pediatric hydrocephalus care.

## Background

Hydrocephalus, characterized by abnormal ventricular enlargement of cerebral ventricles, represents the leading indication for brain surgery in pediatric populations^1^. Accurate quantification of brain and CSF volumes is essential for guiding treatment decisions, enabling clinicians to move beyond subjective visual assessments toward objective, data-driven evaluation. Automated volumetric segmentation further facilitates longitudinal monitoring of brain growth and treatment response^2^.

Clinical management relies on structural imaging modalities such as head ultrasound (HUS) and magnetic resonance imaging (MRI) to monitor ventricular size. Current clinical practice employs proxy measures for ventricular volume, including the frontal and occipital horn ratio (FOHR) for MRI^3^ or ventricular index (VI) for HUS^4^, which are manual linear measurements taken from representative image slices. However, these approaches often introduce subjectivity and fail to accurately capture the complex three-dimensional ventricular morphology present in hydrocephalus. Furthermore, while controlling ventricular expansion remains crucial, promoting healthy brain growth and optimizing long-term neurodevelopmental outcomes constitute the ultimate therapeutic goals for pediatric hydrocephalus management^5–7^.

Comprehensive quantitative assessment of MR brain and cerebrospinal fluid (CSF) volumes requires image segmentation—a labor-intensive process currently limited to research settings or dependent on a new proprietary MR sequence protocol^8,9^. Current state-of-the-art (SOTA) segmentation algorithms rely on intensity-based discrimination between tissue types, assuming consistent intensity distributions between CSF and brain parenchyma. However, the performance of these approaches is suboptimal for challenging pediatric hydrocephalus cases, particularly those with structural brain abnormalities or suboptimal imaging quality^10,11^, limiting their applicability for clinical decision making. Despite the clear clinical need, rapid and accurate automated intracranial segmentation tools suitable for integration into clinical workflows remain notably scarce.

Recent advances in computational capabilities have enabled deep learning (DL) models to demonstrate exceptional performance in complex image processing tasks, particularly segmentation, surpassing conventional intensity-based approaches. The U-Net^12^ architecture stands as a pioneering model in this domain, adept at extracting and utilizing intricate patterns across multiple scales for effective segmentation. However, the sophisticated capabilities of U-Net^12^, derived from its architectural complexity, render its performance highly dependent on training data quality and quantity. This dependency frequently results in overfitting when working with limited datasets, especially when domain-specific knowledge is insufficient. This challenge becomes particularly noticeable in the application to pediatric hydrocephalus. The scarcity of labeled training data combined with the heterogeneity of structural brain abnormalities and variable imaging quality (affected by noise, artifacts, and motion-induced blurring) significantly complicates model training.

To address this critical clinical need, we aim to develop an automated AI-based intracranial segmentation model specifically designed to process clinical MRI scans from infants with hydrocephalus. Our approach integrates advanced deep learning architectures with clinical workflow requirements, emphasizing reliability, speed, and robustness under real-world conditions. We conducted comprehensive validation using 167 clinical MRI scans from infants with hydrocephalus treated at Boston Children’s Hospital, representing the full spectrum of etiologies, imaging qualities, and clinical complexities encountered in tertiary pediatric neurosurgical practice. The technical validation demonstrates superior performance compared to SOTA methods while achieving processing speeds compatible with routine clinical workflows. The study establishes the technical foundation for clinical implementation of AI-based volumetric assessment in pediatric hydrocephalus management, with potential to significantly improve clinical decision-making consistency, workflow efficiency, and patient care quality.

## Methods

### Dataset

Clinical MRI studies from patients undergoing endoscopic third ventriculostomy combined with choroid plexus cauterization (ETV/CPC) for hydrocephalus treatment at Boston Children’s Hospital from 2008 to 2021 were collected retrospectively for the study. Inclusion criteria were patients less than one year old at the time of treatment and at least one preoperative axial T2 weighted (T2w) brain MRI scan available for analysis. Exclusion criteria were hydrocephalus secondary to encephalocele and incomplete intracranial volume coverage precluding accurate intracranial segmentation. One axial T2w brain MRI scan within two weeks before surgery was included per patient. The study received ethical approval from the institutional review board at Boston Children’s Hospital.

The dataset consisted of 167 axial T2w scans. Males accounted for 97 (58%) of patients and the median (IQR) corrected age at scan was 34 (3 – 124) days. Etiologies included aqueductal stenosis (n = 37), dandy-walker complex (n = 23), myelomeningocele (n = 51), post-hemorrhagic hydrocephalus (n = 52), and post-infectious hydrocephalus (n = 4). The dataset was randomly divided into 101 (60%) for training, 33 (20%) for validation, and 33 (20%) for hold-out testing.

### MRI acquisition

MRI scans were obtained from one of the following clinical scanners: Siemens Healthineers, GE healthcare, and Philips Healthcare. MRI was performed at 1.5T or 3T. The T2-scan protocols included fast-spin echo (FSE) T2, T2 blade, T2 propeller, and fast recovery FSE (FRFSE) T2, with a mean slice thickness of 3.2 mm (range 1 - 5). Scans were retrieved via the ChRIS Research Integration System (ChRIS)^13,14^.

### Ground Truth Image Segmentation

The creation of ground-truth segmentation employed an active learning scheme, involving several cycles of manual annotation, model training, and inference. This strategy significantly reduces the time spent on manual segmentation. More specifically, the process began with annotating a diverse selection of scans using ITK SNAP^15^ by trained staff (M.H.Y. and A.S.L.) supervised by a senior pediatric neuroradiologist (P.E.G.). These annotated scans then were used to train the AI segmentation model. Once trained, the model was applied to the remaining unlabeled scans. The results of these inferences were then visually inspected and the scans with the poorest results were manually corrected. After being revised, the segmentations were included in the next training cycle. This iterative process was repeated until no significant improvements were observed in the model. Three classes were created - CSF, brain parenchyma, and background. Brain parenchymal segmentation started at the first superior axial slice where the brain parenchyma was visible and continued to the level of the foramen magnum. Background is defined as any region that is not either CSF or brain parenchyma.

### AI-based Brain Parenchyma and Cerebrospinal Fluid Volumetry Segmentation Model

In Fig. 1, the pipeline of the brain parenchyma and CSF volume estimation model from DICOM raw files was illustrated, which included four steps: information extraction, image pre-processing, AI-based image processing, and volume calculation.

**Fig. 1.**
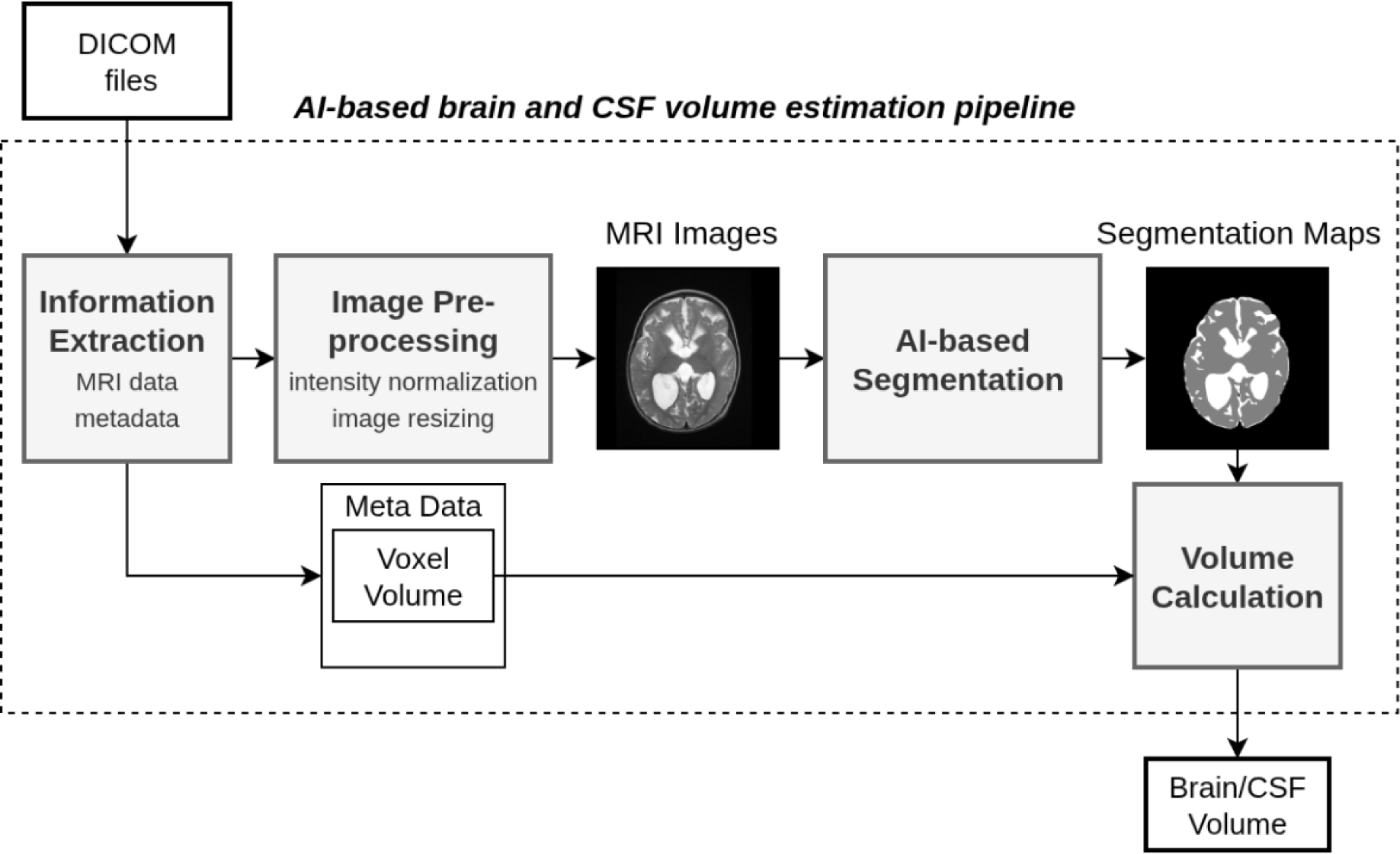
The pipeline of AI-based brain and CSF volume estimation. This figure illustrates the pipeline for AI-based brain and CSF volume estimation. The pipeline takes a patient’s DICOM files as input and outputs the estimated brain and CSF volumes. It consists of four main components: information extraction, image preprocessing, AI-based segmentation, and volume calculation.

As the initial step, raw MRI data along with metadata, which included patient information and scan details, were extracted from DICOM files. The MRI data was then stored and visually inspected to ensure accuracy.

Next, the MRI images are pre-processed, involving intensity normalization and image shape standardization. For intensity normalization, pixel intensities were scaled such that the 99.7^th^ percentile intensity became the maximum value. Intensities exceeding this threshold were capped. Each pixel’s intensity was then divided by this maximum, scaling all images to a range from 0 to 1. Additionally, all MRI images were standardized to a size of 512×512 during training, using either cropping or zero-padding (only two scans in our dataset were slightly larger than this size). During inference, each image is resized to the nearest multiple of 16—using cropping or padding as needed—to ensure compatibility with the encoder-decoder architecture of the network. For example, images sized 412×412 and 418×418 would both be resized to 416×416 through padding and cropping, respectively.

The preprocessed MRI images are sent to an AI-based segmentation model (Fig. 2). A U-Net^12^ is leveraged as the backbone architecture. This structure is renowned for its efficiency in leveraging both local and global semantic information for pixel-level prediction. In the encoding path, the model extracts higher-level features that represent semantic information. These features are then progressively decoded through the decoding path to reconstruct the target output. The skip connection strategy, where low-level intermediate features from the encoding path were combined with high-level features in the decoding path, allowed the network to utilize a comprehensive blend of information. Additionally, to address the challenging diversity presented by clinical MRI scans in patients with hydrocephalus, the standard convolutional blocks in the original U-Net^12^ were replaced by the more powerful Dense blocks of DenseNet^16^. Dense blocks are characterized by their densely connected layers, where each layer receives feature maps concatenated from all previous layers within the block. Therefore, the final output of the entire Dense Block was a rich, multi-scale feature representation. This output captures both low-level and high-level features, making it highly informative for further processing layers in the network. In our model, each dense block contains 4 densely connected layers (Fig. 3). Subsequent to each Dense Block, a Transition Block was utilized to compact the feature maps, consisting of a convolutional layer followed by 2×2 average pooling. This reduction in dimensions was complemented by an expansion in the field of view of the following convolutional layers, significantly enhancing the network’s capability to capture broader, more global features. The final parameterized layer was a 1×1 convolution layer that transforms the deeply aggregated features into voxel-wise class scores for non-brain, CSF, and brain parenchyma. At last, applying a softmax function to the scores of each pixel produces estimated probabilities for each class at every pixel, which is formulated as:

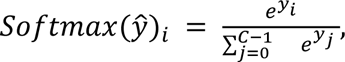

**Fig. 2.**
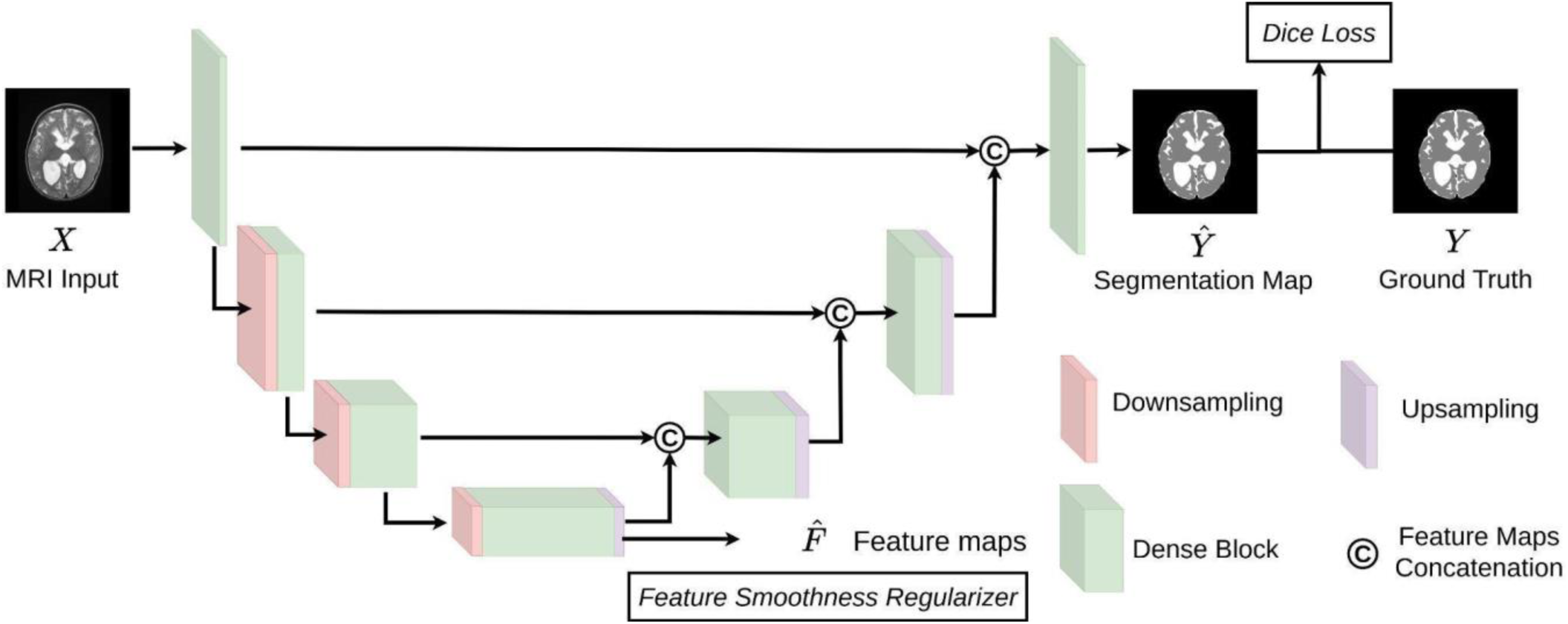
Diagram of our AI-based MRI segmentation model, following a U-Net^12^ architecture with skip connections that bridge encoder and decoder layers, allowing detailed spatial information from early layers to guide accurate reconstruction in later stages.

**Fig. 3.**
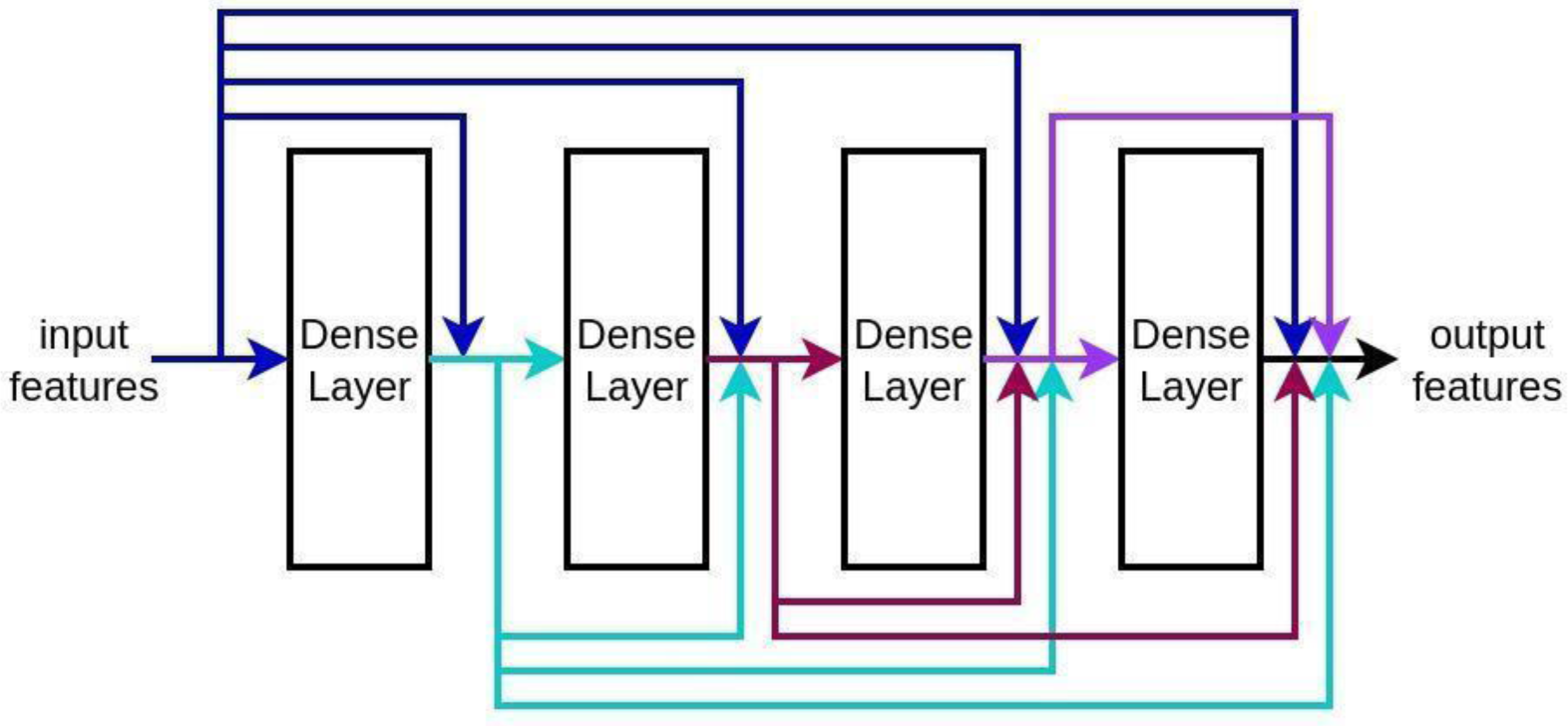
Illustration of features connections inside the Dense Block. The output of a Dense Block was a concatenation of all the feature maps produced by its constituent layers.

where *ŷ* represents the vector of the pixel’s estimated probabilities. The class with highest estimated probability is outputted as the prediction for each voxel. Details about model training are described in the section of Model Training.

In the final step, the brain and CSF volumes are estimated by multiplying the number of voxels predicted as brain and CSF regions by the voxel volume obtained from the DICOM metadata. Here, each pixel from the 2D image slice is treated as an approximate voxel to facilitate volumetric computation.

### AI Model Training

We utilized Dice loss as the main objective function, derived from the Dice coefficient (also known as the Sørensen-Dice coefficient^17^), a commonly used metric for assessing the overlap in segmentation tasks. Dice coefficient is formulated as:

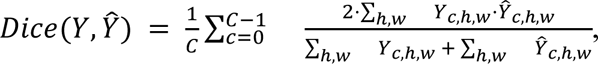

where *Y* is the ground-truth label map and *Ŷ* is the output of the segmentation model. Both consist of C=3 maps (non-brain, CSF, and brain parenchyma), each with a shape of H×W. Therefore, Dice loss is defined as:

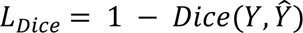

A lower Dice Loss indicated a higher degree of similarity between the predicted segmentation and the ground truth. By minimizing this objective function, the model maximized the overlap between its predicted output and ground-truth labels.

In addition to the standard Dice loss, we introduced a feature smoothness (FS) loss *L*_*fs*_, which acted as a regularizer to the main loss term:

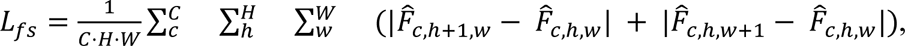

where *F̂* are the feature maps output from the encoder. This regularization term quantified the total spatial variation in the feature maps. The rationale behind this regularizer was twofold: first, segmenting CSF and brain parenchyma depended more on broader contextual information than on local details; second, artifacts and noise, which were prevalent in clinical MRI scans, should not dominate the model’s focus. By appropriately moderating this term, the model was penalized for overfitting fine-grained details, artifacts, or noise. Consequently, the model was encouraged to seek simpler and more generalizable features during training, a strategy that was particularly advantageous when limited training data were available.

In summary, the overall objective function was as follows: *L* = *L*_*Dice*_ + *α L*_*fs*_, where *α* is a hyper-parameter weighing the regularizer which is selected via 5-fold cross-validation^18^.

By effectively combining and balancing the main Dice loss with the FS regularizer, the overall objective function directed the model to learn the most discriminative and generalizable features, thereby enhancing the accuracy of segmentation tasks.

Data Augmentation: To enhance model robustness and mitigate overfitting, various data augmentation techniques were dynamically implemented during training. These included rotations, flipping (horizontal and vertical), resizing, and addition of Gaussian noise. These augmentations were applied on the fly, ensuring a diverse range of features for the model to learn from.

Pre-training: Self-supervised learning, including the use of autoencoders for pretraining, leveraged unlabeled data to learn useful representations before training, which was particularly valuable in medical imaging where labeled data can be scarce and expensive to obtain^19,20^. During the pre-training phase of our model, an autoencoder was formed, consisting of an encoder and a decoder with the final layer transformed as a single channel output. The autoencoder was trained on unannotated 380 T2 MRI scans of hydrocephalus patients from Boston Children’s Hospital to reconstruct the original T2 image directly from the input. The L2 norm loss was used as the objective function, and the learning rate was set to 0.0001 over 300 epochs. After pre-training, only the encoder’s parameters were retained and transferred as initialization for the encoder of our model. The decoder was initialized using He’s initialization^21^. This self-supervised pre-training method benefited the training process in that the encoder was already accustomed to the T2 MRI scans from BCH, leading to lower training loss and faster convergence.

Setting for Training and Inference: The model was trained with a batch size of 8. The Adam optimizer was adopted with a weight decay set to 10^−6^, optimizing the balance between learning speed and regularization to avoid overfitting. *α* in the overall objective function is set to 0.01. The GPU used for training and inference was NVIDIA A30. The CPU was AMD Ryzen 9 7950X Processor 16-Core. The model was trained for 600 epochs. The learning rate was initialized at 0.0001 and scheduled to reduce to 20% of its previous value at epoch milestones of 200 and 400 to ensure gradual convergence.

### Volume Calculation

The number of voxels assigned to each tissue class in the AI segmentation output is multiplied by the voxel volume derived from the MRI metadata to compute the corresponding brain or CSF volume.

### Evaluation Metric

During the evaluation phase, we assessed the model’s performance on the test data using the Dice coefficient, the Absolute Relative Volume Error (ARVE) (Eqn. 1) and Relative Volume Error (RVE) (Eqn. 2), all computed at the scan level. We report both the mean and standard deviation for each evaluation metric. The Dice coefficient measures the spatial overlap between the predicted and ground truth segmentations, indicating the accuracy of the segmentation boundaries. The Absolute Relative Volume Error (ARVE) quantifies the absolute deviation between the predicted and ground truth volumes, normalized by the ground truth volume. The Relative Volume Error (RVE) serves as an auxiliary metric to assess whether the model’s volume estimates exhibit systematic bias—i.e., consistent overestimation or underestimation. These metrics are particularly important because they directly reflect the model’s accuracy and reliability in producing clinically meaningful volume measurements.

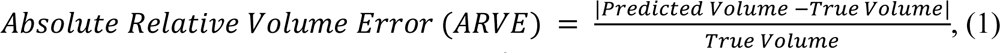

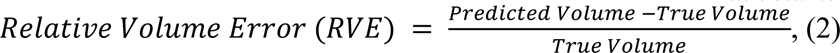

### Statistical analysis

We compare model performance by Dice score, ARVE and RVE on the scan-level. Since all three variables from three methods are deviated from normality (Shapiro–Wilk^22^ p-values < 0.05), the Kruskal-Wallis test^23^ was applied for overall comparison across the three methods, followed by the Mann–Whitney U test^24^ with Bonferroni correction for multiple comparisons.

## Results

### Quantitative Results

We benchmark against two prominent methods: the FSL FAST algorithm^25^ and Grimm *et al.* ’s method^26^. The FSL FAST (FMRIB’s Automated Segmentation Tool)^25^ algorithm is a well-established method in MRI T2 segmentation, utilizing intensity-based statistical modeling to differentiate tissue types. Grimm *et al. ’s* method^26^ is a deep learning approach, which adopted an encoder-decoder architecture^27^. We applied FSL^25^, Grimm *et al. ’s* method^26^ (trained on our dataset), and our model on the hold-out test set. The mean and standard deviation of Dice coefficients across scans are summarized in Table 1. Results of ARVE and RVE are summarized in Table 2.

**Table 1.** Dice score from our approach and SOTA methods. Mean and standard deviation are reported.

| Approach | CSF | Brain parenchyma |
| --- | --- | --- |
|  | Dice score | Dice score |
| Our model | 95.7%±2.3% | 96.4%±2.5% |
| FSL<sup>25</sup> | 85.0%±10.5% | 77.9%±9.1% |
| Grimm *et al.*<sup>26</sup> | 90.4%±3.5% | 89.7%±4.5% |

**Table 2.** ARVE and RVE from our approach and SOTA methods. Mean and standard deviation are reported.

| Approach | CSF |  | Brain parenchyma |  |
| --- | --- | --- | --- | --- |
|  | ARVE | RVE | ARVE | RVE |
| Our model | 2.6% ± 4.7% | 0.03% ± 5.4% | 1.8% ± 2.2% | -1.2% ± 2.6% |
| FSL<sup>25</sup> | 22.9%±23.3% | -3.5%±32.5% | 35.7%±25.3% | 34.6% ±26.7% |
| Grimm *et al.*<sup>26</sup> | 5.7% ± 5.6% | -1.6% ± 3.5% | 9.6% ±12.1% | 7.8% ± 7.4% |

The quantitative results are presented in Tables 1 and 2. Our model achieves Dice scores of 95.7% for CSF and 96.4% for brain parenchyma, surpassing state-of-the-art (SOTA) methods and demonstrating high segmentation accuracy. For both brain parenchymal and CSF volumes, the Dice scores achieved by our model are significantly higher than those of FSL^25^ (p-value < 0.0001) and Grimm *et al.* ’s method^26^ (p-value < 0.05). Box-and-whisker plots are shown in Fig. 4. In addition, the ARVEs of our method are significantly lower than those of existing approaches, with ARVE values of 2.6% for CSF and 1.8% for brain, outperforming SOTA methods for both brain (p-value<0.005) and CSF (p-value<0.005). The RVE for CSF is 0.03%, indicating no apparent estimation bias, while the RVE for the brain is -1.2%, suggesting a slight tendency toward systematic underestimation. This performance enhancement underscored our method’s advanced capability to handle diverse and challenging segmentation scenarios for hydrocephalus MRI, providing robust, accurate, and efficient segmentation outputs essential for clinical application.

**Fig. 4.**
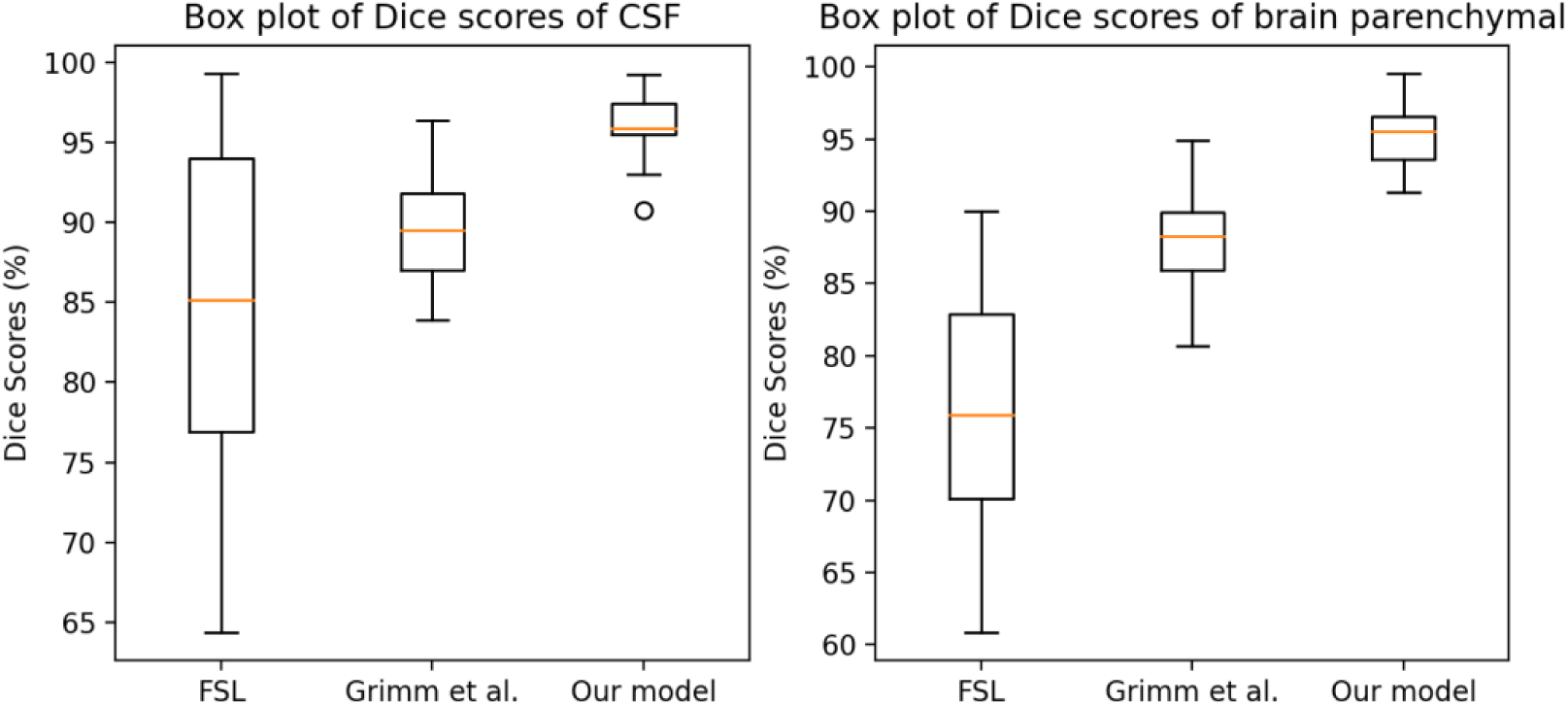
Box plots of Dice scores of CSF (left) and brain parenchyma (right) from SOTA methods and our model. Box represents 50% of data.

Results of etiologies: We report the Dice score for each etiology in Table 3. The consistency in Dice scores across different etiologies demonstrated that the model performance is robust and consistent regardless of the underlying cause of hydrocephalus.

**Table 3.** Dice scores of five etiologies by our model.

| Etiologies | CSF | Brain parenchyma |
| --- | --- | --- |
| Aqueductal stenosis | 95.5% ± 2.4% | 96.4% ± 2.4% |
| Dandy-Walker | 96.4% ± 2.7% | 95.8%± 3.1% |
| Myelomeningocele | 96.7% ± 3.4% | 97.3% ± 2.4% |
| Post-hemorrhagic | 95.9% ± 2.5% | 96.7%± 2.5% |
| Post-infectious | 95.2% ± 2.3% | 96.4%± 3.5% |

Effect of FS regularizer: In Table 4, we present a comparison between the model with and without the FS regularizer. The results demonstrate that incorporating the regularizer leads to a notable improvement (p-value<0.05 for CSF) in performance, particularly for the CSF segmentation. Specifically, the mean Dice score increases by 2%, while the standard deviation is reduced by 1%, indicating both enhanced accuracy and greater consistency across samples.

**Table 4.** Dice score comparison of models trained with and without the FS regularizer.

| Model | CSF | Brain parenchyma |
| --- | --- | --- |
| Our model without FS regularizer | 93.7% ± 3.4% | 95.7% ± 2.8% |
| Our model with FS regularizer | 95.7% ± 2.3% | 96.4% ± 2.5% |

Results of various resolutions: In Fig. 5, we present the average Dice scores of our model’s performance on the test dataset across different MRI resolutions. For each resolution level, scores from multiple scans were averaged to represent the overall performance at that resolution. As shown in the figure, the model demonstrates consistent performance across varying resolutions, primarily due to the inclusion of diverse resolution levels in the training set and the use of data augmentation.

**Fig. 5.**
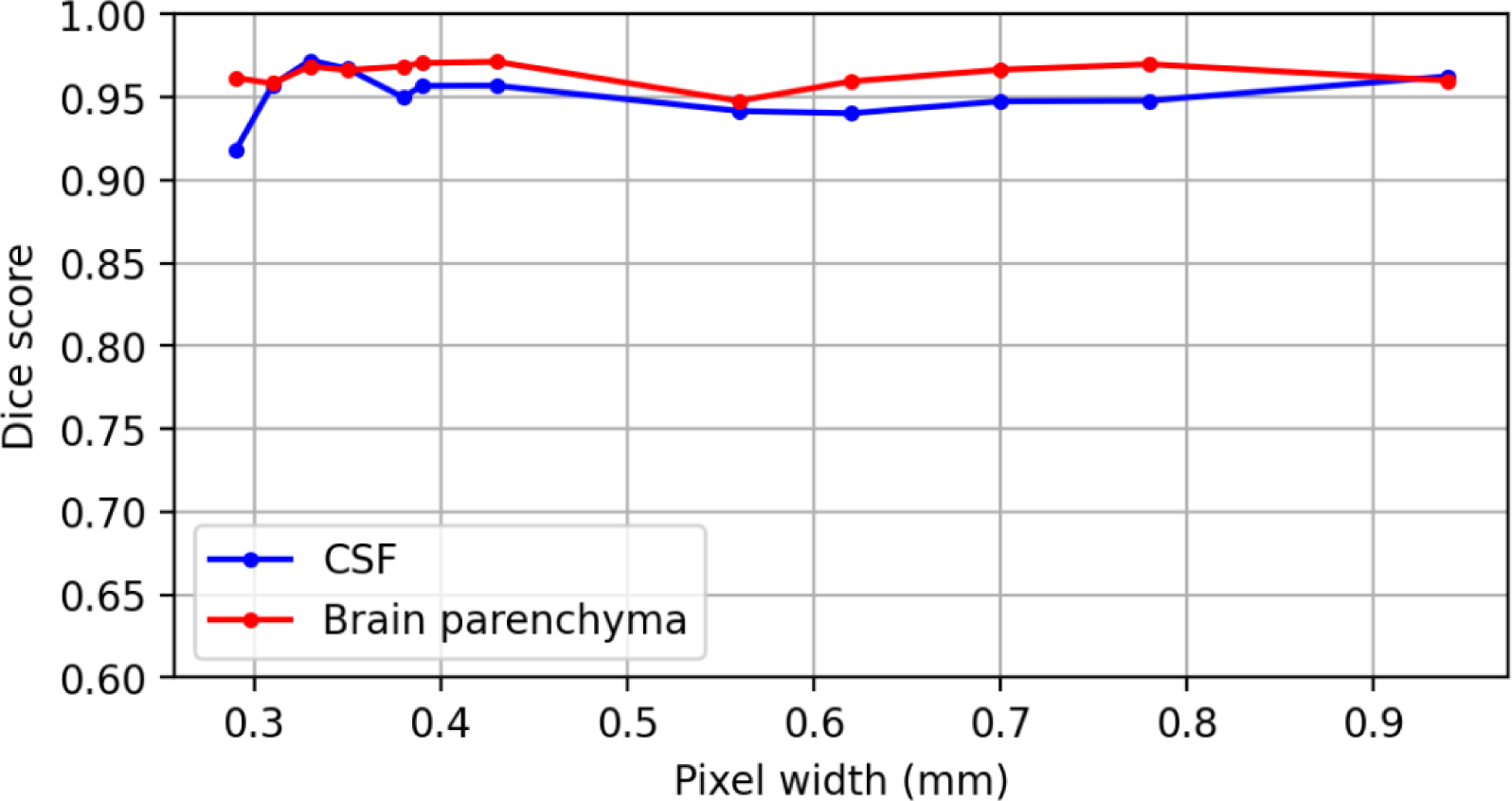
Average Dice scores of our model’s results on the test dataset across different MRI resolutions (with higher resolution indicating smaller pixel widths). The plot shows that our model consistently achieves high performance across varying MRI resolutions.

### Processing Time

Furthermore, compared to FSL^25^, our AI-based approach obtained much faster processing times. Processing a complete scan takes 0.8 seconds with our model using a NVIDIA A30 GPU and 20 seconds using a CPU (AMD Ryzen 9 7950X Processor 16-Core), whereas FSL^25^ requires approximately 2 minutes to process one scan using a CPU with the same settings.

### Representative examples Comparison with SOTAs

Fig. 6 exhibited the underlying MRI scans and ground-truth segmentation along with the segmentation results from our method, FSL^25^, and Grimm *et al*’s^26^. It is evident that our approach effectively excluded eyeballs, skulls, and other non-brain objects from the segmentation. Furthermore, in the examples of inhomogeneous imaging intensities due to CSF flow shown in the right panel of the figure, our method continued to produce reasonable outputs, showcasing its successful application of global features extracted through encoder and enhanced by FS regularizer.

**Fig. 6.**
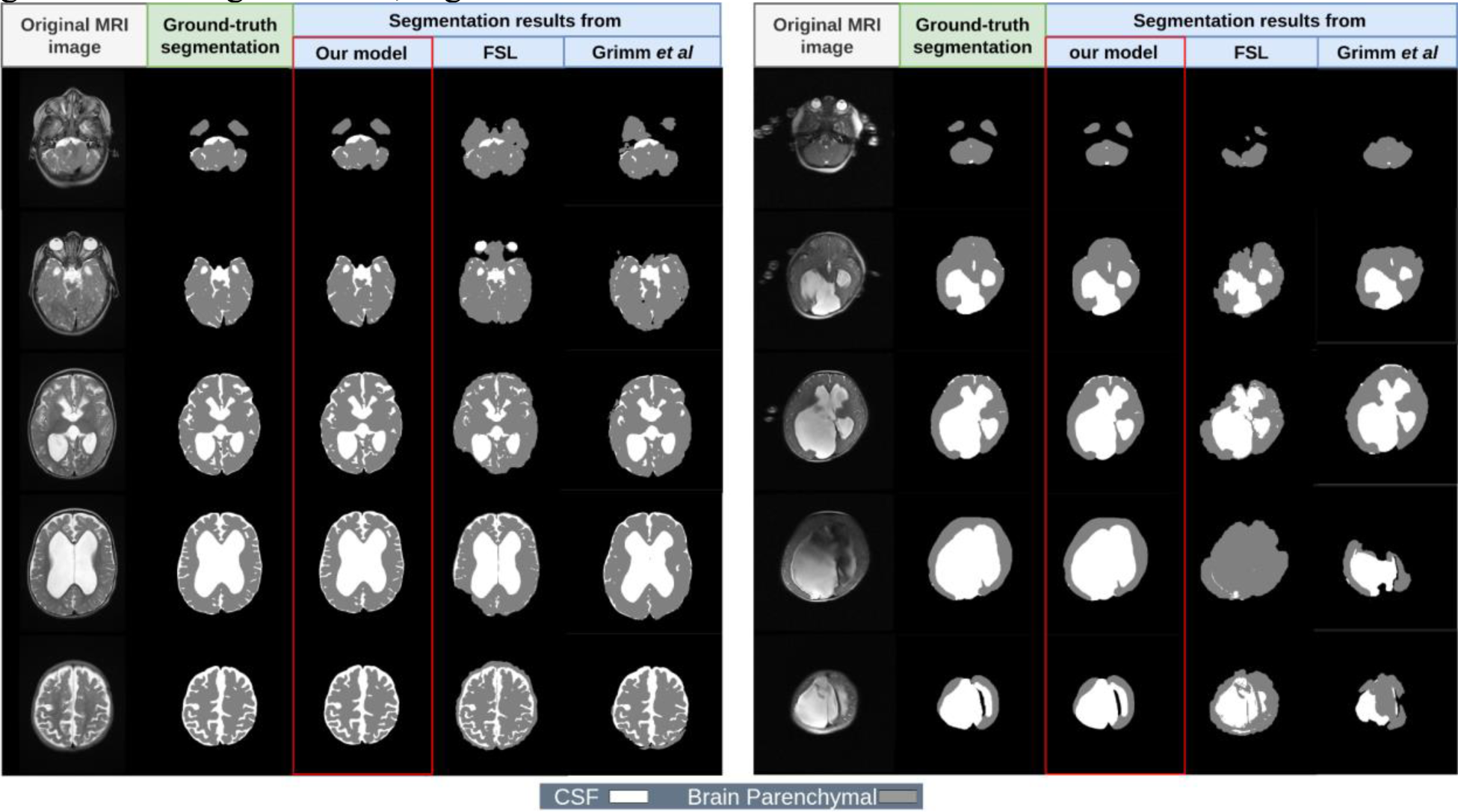
Segmentation examples of clinical scans. From left to right: original T2-weighted images, ground truth segmentation, segmentation results of our method and FSL^25^.

### Segmentation Results on Challenging Clinical Scans

Overall, our AI model demonstrates remarkable robustness in handling the diverse artifacts and noise commonly encountered in pediatric MRI.

### Motion Artifacts

In the top examples of Fig. 7, despite significant motion-induced blurring and distortion that degrades image quality, our model maintains precise segmentation boundaries between CSF and brain tissue, accurately following the ground-truth segmentation. The ability to handle motion artifacts is critically important given that motion artifacts are extremely common in pediatric imaging due to the difficulty of keeping infants still during scanning.

**Fig. 7.**
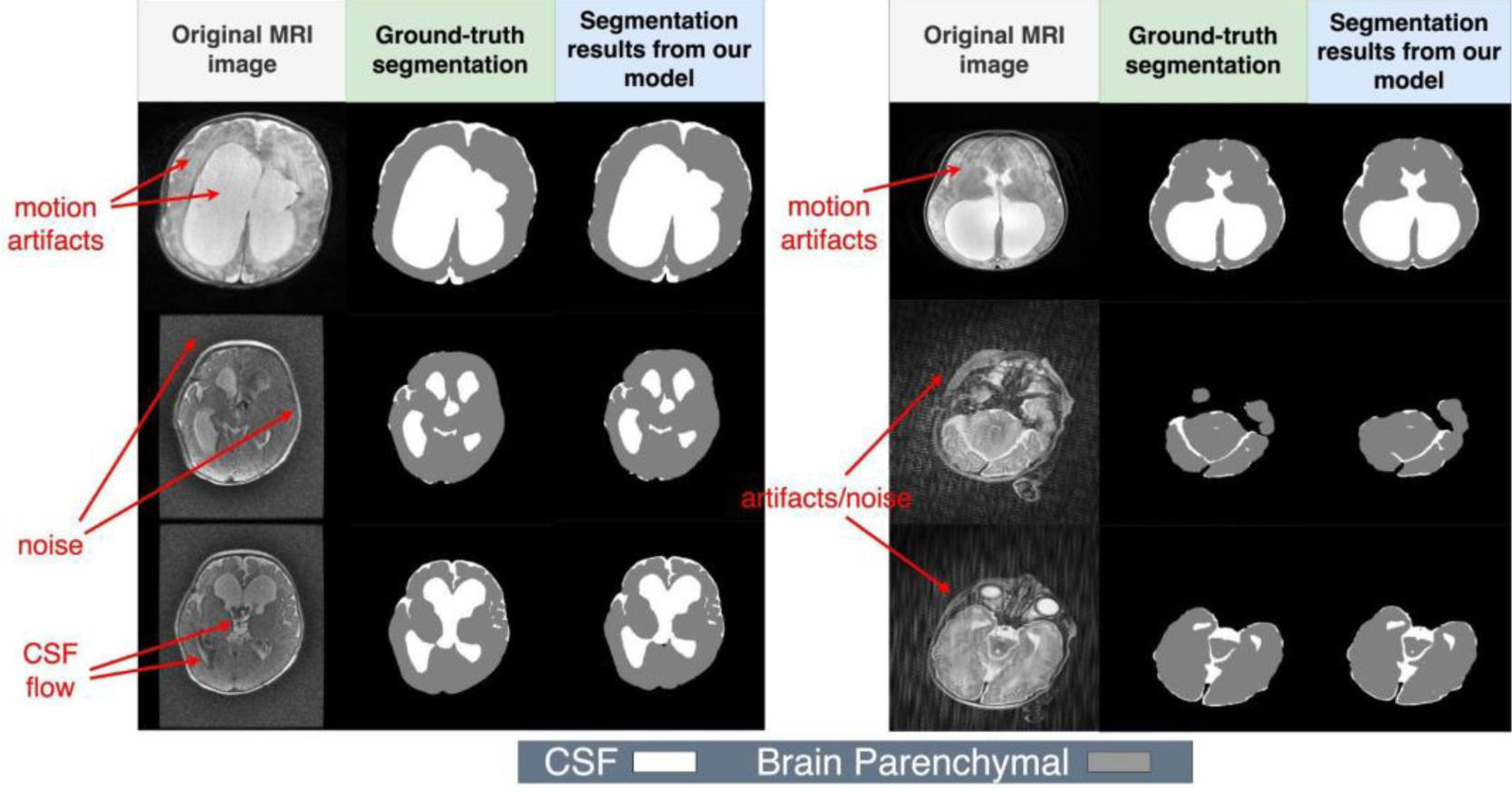
Results of challenging scenarios where noise and artifacts are present in the scans.

### Image Noise

The middle-left example of Fig. 7 shows substantial noise corruption that creates a grainy, low-contrast appearance throughout the image. Despite this degraded signal quality, our model successfully generates consistent segmentation masks that closely match the ground truth, demonstrating its ability to extract meaningful patterns of CSF and brain from highly corrupted signals.

### CSF Flow Artifacts

The bottom-left case of Fig. 7 illustrates CSF flow-related intensity variations that create irregular signal patterns within the ventricular spaces. These flow artifacts can confuse intensity-based segmentation methods, but our AI model correctly identifies these regions as CSF while maintaining accurate boundaries with surrounding brain tissue.

### Combined Artifacts/Noise

The right panel of Fig. 7 examples show complex combinations of artifacts and noise that create particularly challenging segmentation scenarios. Even under these challenging conditions, the AI model produces segmentations that are nearly indistinguishable from ground truth,.

## Discussion

This study addresses a critical gap in pediatric hydrocephalus management by developing an AI-based segmentation model that achieves clinically viable accuracy for automated volumetric analysis. Our model demonstrates substantial improvements over existing methods, with Dice scores of 95.7% for CSF and 96.4% for brain parenchyma, while maintaining processing speeds compatible with routine clinical workflows (0.8 seconds per scan). These results represent meaningful progress toward making quantitative volumetric assessment practical for clinical use. This advancement enables routine volumetric monitoring that can support treatment monitoring and longitudinal assessment of neurodevelopmental outcomes.

Existing studies on brain segmentation primarily rely on intensity-based methods or atlas-based features^28–32^. Most current automated tools for intracranial segmentation are designed for small cohorts of healthy infants and often suffer from long processing times, limiting their clinical applicability^33–40^. More critically, pediatric hydrocephalus presents unique technical challenges that distinguish it from general neuroimaging segmentation tasks. The pathophysiology fundamentally alters normal brain anatomy, with excessive CSF accumulation creating substantial shifts in intensity distributions compared to healthy infants. Unlike adult brains where tissue boundaries are well-defined, hydrocephalus cases exhibit highly variable ventricular morphology, compressed brain parenchyma, and altered CSF flow patterns that confound standard intensity-based segmentation approaches. The heterogeneity of hydrocephalus etiologies—including aqueductal stenosis, post-hemorrhagic hydrocephalus, myelomeningocele, and dandy-walker complex—introduces additional anatomical variations that challenge automated algorithms. Furthermore, the rapidly changing brain development in infants creates challenges for segmentation models, as normal anatomical references vary significantly across age groups. These factors combine to create a technical landscape where conventional segmentation tools, designed primarily for healthy adult brains such as FSL^25^, consistently underperform or fail entirely.

The clinical environment for pediatric hydrocephalus imaging introduces additional complexity beyond the inherent technical challenges. Motion artifacts are common due to the difficulty of keeping infants still during scanning without sedation. Variable imaging protocols across institutions create inconsistent data quality with differences in slice thickness (1-5mm in our dataset), field strengths (1.5T vs 3T), and sequence types (FSE T2, T2 blade, T2 propeller, FRFSE T2) that standard algorithms may struggle to accommodate. Clinical scans frequently contain noise, CSF flow artifacts, and intensity inhomogeneities that are absent from carefully controlled research datasets^41^. Another critical challenge is the limited availability of annotated data^42^. Since infant hydrocephalus is usually treated at specialized children’s hospitals, collecting large, well-annotated datasets for training is difficult, further complicating the development of robust automated solutions.

In recent years, pioneering studies have applied machine learning techniques to hydrocephalus segmentation, achieving promising results. Notably, Cherukuri *et al.* developed a learning-based CT segmentation algorithm that accurately delineates the brain, CSF, and subdural regions, showing strong performance in clinical applications^43,44^. Building on this foundation, Yu *et al.*^45^ developed domain-enriched attention mechanisms for infection diagnosis in hydrocephalus CT images, demonstrating that incorporating domain-specific knowledge into deep learning architectures significantly improves performance on challenging clinical datasets. Moreover, numerous studies have employed deep learning, particularly U-Net–based models, on segmentation tasks for pediatric patients. Grimm et al.^26^ leveraged 2D U-Net^12^ for pediatric hydrocephalus. Largent et al.^46^ advanced the field with 3D Bayesian U-Net approaches achieving 95% Dice scores in preterm infants with post-hemorrhagic hydrocephalus; however, their reliance on 3D volumetric data limits clinical applicability since routine practice predominantly uses 2D acquisitions due to time constraints and patient motion.

Recent efforts have increasingly focused on improving model robustness. Ren *et al.*^32^ introduced attention-based mechanisms to enhance segmentation consistency, while Qiao *et al.*^47^ proposed spatial guidance strategies to achieve more accurate boundary delineation. Mukherjee *et al.*^48^ further developed a hybrid classification–segmentation framework, representing the first work to segment low-field hydrocephalus MRIs.

Although these methods have achieved notable success in their respective tasks and demonstrated promising clinical results, transferring machine learning models to different datasets remains challenging due to image- or domain-shift issues. General pediatric brain segmentation tools^49,50^ have shown substantial performance declines when applied to hydrocephalus cases, largely because of fundamental anatomical and intensity distribution differences. Another important challenge across these approaches lies in maintaining robustness under varying imaging conditions and hydrocephalus etiologies.

To tackle the unique challenges observed in our patient cohort, we constructed a comprehensive, clinically curated dataset encompassing a wide range of hydrocephalus etiologies and imaging variations representative of tertiary pediatric neurosurgical practice. Using this dataset, we developed an improved AI model specifically optimized for these conditions. Experimental results show that the proposed model surpasses conventional and state-of-the-art methods, demonstrating strong potential for clinical deployment in specialized pediatric hydrocephalus care settings. The achieved Dice scores of 95.7% for CSF and 96.4% for brain parenchyma represent significant advances over existing methods—FSL^25^ achieved only 85.0% and 77.9% respectively, while the state-of-the-art deep learning approach by Grimm *et al.*^26^ reached 90.4% and 89.7%. More critically, our results address the fundamental performance gaps of existing state-of-the-art models when applied to pediatric hydrocephalus highlighted by recent evaluations^51^, where leading models (FastSurfer and QuickNAT) achieved only 0.61 Dice coefficients with 30% complete failure rates in challenging cases.

The robust performance stems from several key innovations that directly target the limitations of existing approaches. The integration of DenseNet^16^ and U-Net^12^ architectures creates rich, multi-scale feature representations that capture both local tissue characteristics and global anatomical context—addressing the challenge of variable ventricular morphology that confounds conventional networks. Unlike the generic architectures used in previous deep learning approaches, our FS regularization mechanism specifically counters the tendency to overfit to artifacts and noise common in clinical scans, promoting focus on anatomically meaningful features essential for hydrocephalus cases.

The quantitative results validate these design choices. Our absolute relative volume errors (ARVE) of 2.6% for CSF and 1.8% for brain parenchyma indicate clinically acceptable accuracy. The relative volume error (RVE) for CSF approximates zero (0.03%), confirming unbiased estimation, while the slight brain parenchyma underestimation (-1.2%) represents a clinically negligible bias (e.g., approximately 5 cm³ underestimation for a brain volume of 500 cm³). Statistical significance testing confirms superior performance over both standard and contemporary approaches. Critically, our approach addresses the generalization challenges that have limited previous methods. The consistency across different hydrocephalus etiologies (Table 3) and imaging resolutions (Fig. 5) demonstrates the robustness essential for broad clinical deployment that previous approaches lacked. Our self-supervised pre-training strategy leverages unlabeled hydrocephalus data to initialize the encoder with domain-specific knowledge, directly addressing the limited training data challenge that has constrained other methods. Most importantly, unlike existing tools that have higher fail rate in challenging cases^45^, our specialized approach achieved 100% success rate across 167 diverse clinical cases, successfully processing challenging scenarios with motion artifacts, noise, and CSF flow abnormalities.

Beyond these quantitative improvements, this advancement significantly streamlines the MRI analysis process. Specifically, when generating a new training dataset, instead of manual segmentation from scratch, our model could generate initial segmentations for manual editing. This approach can reduce time needed for brain segmentation from hours to minutes while still providing highly reliable training datasets for future model development or validation.

While our model demonstrates superior performance on pre-operative MRI scans from infants under one year, several limitations warrant acknowledgment and guide future development priorities. The current dataset focuses on pre-surgical cases and the applicability to post-operative monitoring where CSF-brain parenchyma relationships may change anatomically post-surgically requires further validation. An important consideration involves brain myelination during early development. Progressive myelination over the first 2-3 years substantially alters T2 signal characteristics^52^, potentially affecting segmentation performance. While our training dataset and 380 pre-training scans include infants spanning different myelination stages within the first year, systematic evaluation across the complete developmental timeline (birth through 3 years) is needed to establish generalizability as the brain myelinates and to determine whether age-specific model refinement is required for clinical deployment beyond infancy. The slight systematic underestimation of brain parenchyma volume, though clinically negligible, requires investigation to understand underlying causes. Our training data originates from a single institution, potentially limiting generalizability across different imaging protocols and patient populations. Future work will expand the dataset to include consecutive MRIs across different age groups, treatment stages, and institutions to enhance model robustness and applicability. Incorporating geometric location information could also lead to enhancement of the model’s accuracy, particularly in anatomically complex regions, thus providing more detailed and clinically relevant segmentations.

Our ultimate goal is to integrate our AI-based volumetric assessment into clinical workflows transforming pediatric hydrocephalus management. The sub-second processing capability enables real-time volumetric analysis during routine clinical visits, allowing clinicians to make immediate treatment decisions based on quantitative brain and CSF volume measurements rather than subjective visual assessments. For longitudinal monitoring, automated volumetric tracking will enable precise assessment of brain growth responses to treatment, supporting evidence-based optimization of hydrocephalus management. In addition, the standardization of volumetric assessment across institutions will support collaborative research initiatives and enable large-scale outcome studies accelerating hydrocephalus research.

## Conclusion

This study demonstrates that domain-specific AI approaches can achieve clinically acceptable performance for automated volumetric assessment in pediatric hydrocephalus, addressing important limitations of existing general-purpose tools. Our integration of DenseNet^16^ and U-Net^12^ architectures with FS regularization provides a practical foundation for clinical implementation, achieving accuracy levels and processing speeds suitable for routine use. The demonstrated robustness across different imaging conditions and hydrocephalus etiologies supports the potential for this tool to improve consistency and efficiency in pediatric neurosurgical workflows. This work contributes to the growing evidence that specialized medical imaging AI tools, designed with domain-specific knowledge, can better address the unique challenges of pediatric populations and pathological conditions than generic approaches.

## Data Availability

All data produced in the present study are available upon reasonable request to the authors.

## Acknowledgments

The authors would like to thank all the administrative, clinical, and research support staff at Boston Children’s Hospital. Specifically, we thank Luis Enrique and the ChRIS team for IT support; Michael Woglom for the help with MRI brain segmentation.

